# Paired skin and serum proteomics capture distinct immune patterns in early childhood atopic dermatitis

**DOI:** 10.64898/2026.11.26.26368947

**Authors:** Karolina Dumycz, Philipp Gessner, Joanna Nowiszewska, Dominika Ambrożej, Anja Heider, Anna Stelmaszczyk-Emmel, Dominika Nowis, Cezmi Akdis, Marek Kulus, Christoph Messner, Milena Sokolowska, Wojciech Feleszko

**Affiliations:** Department of Pediatric Pneumonology and Allergy, Medical University of Warsaw, Warsaw, Poland; Doctoral School, Medical University of Warsaw, Warsaw, Poland; Swiss Institute of Allergy and Asthma Research (SIAF), University of Zurich, Davos, Switzerland; Precision Proteomics Center, University of Zurich, 7265 Davos, Switzerland; Swiss Institute of Bioinformatics (SIB), 1005 Lausanne, Switzerland; Department of Dermatology, Medical University of Warsaw, Warsaw, Poland; Department of Laboratory Diagnostics and Clinical Immunology of Developmental Age, Medical University of Warsaw; Laboratory of Experimental Medicine, Medical University of Warsaw, Warsaw, Poland; The LOOP Zurich, 8044 Zurich

## Abstract

**Introduction:** Atopic dermatitis (AD) is a heterogeneous disease encompassing multiple immune endotypes, yet clinically informative biomarkers remain limited. In particular, the relationships between local skin and systemic immune biomarkers and food sensitization across the pediatric AD severity spectrum are poorly understood, especially in early childhood.

**Methods:** Skin tape strips (STS) from non-lesional skin and paired sera were analyzed by targeted proteomics in children aged 3 months to <6 years with AD (n=49) and age- and sex-matched healthy controls (n=21). Detailed clinical parameters, disease severity scoring, skin transepidermal water loss and corneometry measurements were recorded, and allergen sensitization was assessed using a multiplex allergen panel. Differential expression was assessed across severity and sensitization subgroups, and unsupervised k-means clustering was derived independently in each compartment.

**Results:** Non-lesional skin showed a dominant innate signature (IL-18, CXCL1, CXCL6, IL-8 (CXCL8), CD40) elevated versus controls even in mild disease, with further amplification in moderate-and-severe disease. Eczema Area and Severity Index (EASI) was associated with serum type-2 markers, led by IL-13, with additional associations for MCP-3 and MCP-4. Children sensitized to foods and co-sensitized to foods and aeroallergens showed upregulation of IL-18, CXCL1, CXCL6, IL-8, and CD40 as compared to healthy controls, with no differences between sensitization subgroups. CXCL6 and IL-1α distinguished specific allergen-sensitized groups (tree nuts, egg, storage proteins) from controls. Egg sensitization was associated with elevated serum IL-8. Unsupervised clustering identified two exploratory proteomic clusters per compartment - skin clusters aligned with severity, and serum clusters aligned with age and co-sensitization.

**Conclusions:** Skin and serum proteomes capture different dimensions of pediatric AD heterogeneity: non-lesional skin showed an innate, severity-graded signature detectable already in mild disease, whereas serum clusters aligned with age and sensitization.

## 1. Introduction

Atopic dermatitis (AD) is among the most common chronic inflammatory skin diseases in children, with a prevalence ranging from 2.7% to 20.1% depending on geographic region (1). Although historically conceptualized as a single, Th2-dominant disease, it is now recognized as a heterogeneous condition encompassing multiple phenotypes and endotypes, shaped by age, ethnicity, disease severity, and sensitization status, with contributions from Th1, Th17, and Th22 pathways alongside the Th2 axis (2). This heterogeneity has direct clinical relevance: although Th2-targeted therapies such as dupilumab are effective for many patients, a substantial subset does not respond adequately, underscoring the need for endotype-based patient stratification to guide individualized treatment beyond a single-pathway model (3).

According to the dual-allergen exposure hypothesis, disruption of the epidermal barrier, whether due to filaggrin mutations, environmental insults, or subclinical inflammation, permits allergen penetration and Langerhans cell-mediated Th2 priming, promoting sensitization, whereas early oral exposure favors tolerance (4). Because AD is characterized by impaired barrier function even in non-lesional skin, it represents a plausible cutaneous route for the initiation of allergen sensitization, consistent with the well-documented co-occurrence of AD with food allergy, asthma, and allergic rhinitis over the course of childhood (5).

Skin tape stripping (STS) has emerged as an optimal method for studying skin immune processes, especially in children (6). Unlike skin biopsy, STS is minimally invasive, non-scarring, and suitable for repeated sampling across all pediatric age groups, while still enabling reliable protein, lipid, and RNA-level characterization of the epidermis (7). Critically, STS can be performed on non-lesional skin, allowing detection of subclinical immune and barrier abnormalities that precede or accompany visible disease activity, a feature of particular relevance in milder AD, where non-lesional skin constitutes the large majority of the cutaneous surface, and in the young pediatric population, where skin biopsy is rarely feasible (8).

Despite growing interest in AD endotyping, most proteomic characterizations to date have focused on moderate-and-severe disease, relied on a single biological compartment, or have not been performed in early childhood, when allergic sensitization patterns are still evolving, and endotypes may be most amenable to modification. Therefore, in the present study, we primarily aimed to characterize <u>p</u>roteomic profiles in a cohort of children aged 3 months to <6 years with AD, using paired STS and serum proteomic profiling, in relation to disease severity. Secondly, we aimed to define the role of allergic sensitization defined by component-resolved diagnostics in AD clustering. We further applied an unsupervised, data-driven clustering approach in each biologic compartment to determine whether distinct proteomic subtypes could be identified independently of pre-specified clinical categories, to contribute to a more precise, mechanistically grounded characterization of AD heterogeneity in early childhood.

## 2. Methods

### 2.1 Study participants and clinical evaluation

This cross-sectional study recruited children at the Medical University of Warsaw, including 49 patients with AD and 21 age and sex-matched healthy controls (HC), aged between 3 months and <6 years. Inclusion criteria for the AD group were clinical diagnosis of AD of any severity based on Hanifin and Rajka criteria (9). Exclusion criteria included: (i) active skin infection, (ii) use of systemic immunosuppressive therapy within 4 weeks prior to clinical evaluation, (iii) use of topical corticosteroids or topical immunomodulators within 1 week, or emollients within 12 hours before STS in the skin site of tape stripping, and (iv) the presence of other immune-mediated systemic diseases (e.g., autoimmune conditions) or (v) current infection with symptoms such as fever, acute cough, diarrhea. The control group consisted of healthy children without a history of skin diseases or atopic conditions by the time of recruitment. Demographic and clinical data were collected via caregiver questionnaires. Disease severity in AD patients was assessed using multiple validated instruments: SCORing Atopic Dermatitis (SCORAD), Eczema Area and Severity Index (EASI), Visual Analog Scale (VAS) for pruritus, Patient-Oriented Eczema Measure (POEM), Infants’ Dermatitis Quality of Life Index (IDQOL, for children <4 years), and the Children’s Dermatology Life Quality Index (CDLQI, for children ≥4 years). Disease severity was classified according to established thresholds: mild AD was defined as oSCORAD <15, SCORAD <25, or EASI ≤7; moderate AD as oSCORAD 15-40, SCORAD 25-50, or EASI >7-21; and severe AD as oSCORAD >40, SCORAD >50, or EASI >21 (10,11). Owing to the limited number of patients with severe disease, moderate and severe cases were combined into a single moderate-and-severe group for all analyses. The study was conducted in accordance with the Declaration of Helsinki and was approved by the relevant institutional bioethics committee (KB/80/2022; KB/164/2022). Written informed consent was collected from the legal guardian.

### 2.2 Skin barrier function assessment

Skin barrier function was evaluated by measuring transepidermal water loss (TEWL) and stratum corneum hydration (corneometry) as described in Supplementary materials.

### 2.3. Skin tape strips (STS) and serum samples collection

Minimally invasive epidermal sampling was performed using the skin tape-stripping (STS) technique. Up to fifteen consecutive D-Squame® Sampling Discs (D102, Clinical & Derm LLC) were applied to non-lesional skin preferably on the volar forearm using a standardized D-Squame® Pressure Instrument (D500, Clinical & Derm LLC) to ensure uniform pressure. After each application, tape-strips were removed using gloved hands and tweezers and placed individually onto D-Squame® Standard Storage Cards (D120, Clinical & Derm LLC). Samples were immediately stored at −80°C until further analysis.

Serum was collected using a standardized protocol. Details concerning sample collection are provided in Supplementary materials.

### 2.4 Allergic sensitization assessment

Allergic sensitization was assessed in all children with AD using the ALEX² multiplex allergen test (Allergy Explorer, MacroArray Diagnostics, Vienna, Austria). It measures specific IgE (sIgE) against a broad panel of allergen extracts and molecular components and gives detailed sensitization profiles. sIgE levels ≥0.3 kUA/L were considered positive. A child was classified as sensitized to a given allergen source if sIgE was positive to its extract and/or to at least one of its molecular components.

### 2.5 Protein extraction from skin tape strips

Protein extraction from stratum corneum was performed using the STS collected as described above. For each participant, four tape strips were used for protein extraction. Details concerning protein extraction protocol are provided in the Supplementary materials.

### 2.6. Targeted proteomic analysis of skin tape strips and serum

Quantitative proteomic profiling was performed on both serum samples and protein extracts obtained from STS using the Olink® Target 96 Inflammation panel (Olink Proteomics, Uppsala, Sweden). Details concerning proteomic analysis are provided in Supplementary materials.

### 2.7. Statistical Analysis

All statistical analyses were performed using R (version 4.3.3). Details concerning statistical analysis are provided in Supplementary materials.

## 3. Results

### 3.1 Study cohort

The study cohort comprised 70 children: 33 with mild, 16 with moderate-and-severe AD (based on EASI), and 21 HC (Figure S3). The three groups were comparable with respect to age and sex distribution (Table 1). Lesional skin barrier dysfunction and systemic immune activation, reflected by elevated eosinophil counts, total IgE, CCL17/TARC, and IL-22 levels, were significantly higher in the moderate-and-severe AD group compared to the mild AD group (Table 1). Both groups differed significantly from HC in the majority of measured parameters. Detailed clinical, demographic and sensitization profile characteristics of all participants are provided in Table 1 and Supplementary Tables S1–S3.

**Table 1.** Clinical and demographic characteristics of study participants stratified by EASI-based severity group.

| **Variable** | **AD Mild (n=33)** | **AD ModSev (n=16)** | **HC (n=21)** | **p-value** |
|---|---|---|---|---|
| Sex, female, n (%) | 14 (42.4%) | 8 (50.0%) | 6 (28.6%) | 0.761 |
| Age (years) | 2.74 (1.46-4.28) | 2.73 (2.10-3.55) | 2.07 (1.36-3.23) | 0.678 |
| EASI | 2.00 (1.00-2.90) | 11.60 (9.25-17.15) | 0 | <0.001<sup>\$&</sup> |
| Objective SCORAD (oSCORAD) | 14.20 (7.72-15.48) | 34.67 (30.62-36.87) | 0 | <0.001<sup>\$&</sup> |
| SCORAD (SCORAD) | 17.24 (13.78-26.86) | 47.05 (43.91-48.88) | 0 | <0.001<sup>\$&</sup> |
| Pruritus VAS (0-10) | 4.00 (2.00-5.00) | 7.00 (5.00-7.00) | 0 | <0.001<sup>\$&</sup> |
| Sleeplessness VAS (0-10) | 3.00 (0.00-4.00) | 5.50 (4.75-7.00) | 0 | <0.001<sup>\$&</sup> |
| IDQOL / CDLQI | 5.00 (4.00-8.00) | 14.00 (7.25-16.25) | 0 | 0.001<sup>\$&</sup> |
| POEM | 8.00 (4.00-13.00) | 16.50 (12.75-22.00) | 0 | 0.001<sup>\$&</sup> |
| TEWL non-lesional (g/m²/h) | 16.23 (13.53-19.43) | 16.08 (14.98-21.16) | 13.75 (11.44-14.90) | 0.424<sup>\$&</sup> |
| TEWL lesional (g/m²/h) | 31.66 (27.97-37.39) | 43.53 (38.88-46.84) | N/A | 0.002 |
| Corneometry non-lesional (AU) | 45.74 (37.28-48.77) | 40.81 (32.31-45.45) | 53.13 (46.83-60.20) | 0.163<sup>\$&</sup> |
| Corneometry lesional (AU) | 27.31 (20.94-32.35) | 18.01 (13.97-22.44) | N/A | 0.002 |
| Eosinophils (×10³/μL) | 0.37 (0.23-0.57) | 0.62 (0.48-0.94) | N/A | 0.001 |
| Total IgE (kU/L) | 108.00 (29.90-167.00) | 198.00 (105.15-1420.00) | N/A | 0.016 |
| Vitamin D3 (nmol/L) | 37.20 (29.10-43.90) | 32.50 (28.80-45.00) | N/A | 0.831 |
| CCL17/TARC (pg/mL) | 891.45 (559.08-1378.30) | 1692.42 (978.17-1963.46) | N/A | 0.011 |
| IL-22 (pg/mL) | 17.82 (6.86-30.79) | 102.01 (41.96-128.45) | N/A | <0.001 |

Data are presented as median (IQR) for continuous variables and n (%) for categorical variables. Demographics and non-lesional skin barrier parameters were compared across two groups using Mann-Whitney U test and all three groups using the Kruskal-Wallis test or Chi-squared test. HC data were not collected for AD-specific parameters (indicated as N/A). Presented p-values reflect comparison between AD Mild and AD ModSev. \$ symbol reflects statistically significant difference between HC and Mild AD and & symbol reflects statistically significant difference between HC and ModSev AD. Severity grouping: EASI ≤7 = Mild; EASI >7 = Moderate-and-Severe (ModSev). EASI: Eczema Area and Severity Index. oSCORAD: objective SCORAD. SCORAD: total SCORAD. VAS: visual analogue scale. IDQOL: Infants' Dermatitis Quality of Life index. CDLQI: Children's Dermatology Life Quality Index. POEM: Patient-Oriented Eczema Measure. TEWL: transepidermal water loss. AU: arbitrary units. HC: healthy controls. N/A: data not collected for this group.

### 3.2 Proteomic signatures of non-lesional skin reflect AD severity

The STS proteomic analysis included 67 participants (46 AD, 21 HC). Out of 46 AD subjects with available STS samples, one was excluded for failure to pass assay internal quality control, and one was removed as a proteomic outlier (based on Z-score >3 or <−3 threshold for at least 50% of proteins included in final analysis). Of 92 proteins on the panel, 24 met detection thresholds and were retained for analysis.

Differential expression analysis, adjusted for age and sex, identified 15 proteins differentially expressed between AD and HC. Twelve were upregulated in AD, with the largest effect sizes for IL-18, CXCL1, IL-8, CXCL6, CD40 (all FDR < 0.001), CCL20 and MCP-4 (all FDR < 0.01, Figure 1A, Table S5); DNER, ADA and IL-1α were downregulated (Figure 1A, Table S5). Interestingly, we identified inverse associations between multiple inflammatory proteins and non-lesional skin barrier parameters. Most notably we found that the lower IL-1α expression was associated with higher non-lesional TEWL (ρ = −0.551, p < 0.001, Figure 1B, Table S6), and that higher IL-18, MCP-4, HGF and CDCP1 associated with lower non-lesional corneometry (ρ = −0.476 to −0.404, p<0.05, Table S6), suggesting that these skin inflammatory biomarkers to some extent reflect barrier impairment in non-lesional skin. Next, severity-stratified comparisons against HC, performed across three independent severity scales (EASI, oSCORAD, SCORAD), yielded a consistent core upregulated signature of IL-18, CXCL1, IL-8, CXCL6, and CD40 in both mild and moderate-and-severe AD (Figure 1C, Tables S7-S9). The moderate-and-severe group showed a stronger inflammatory response, with additional upregulation of CCL20, MCP-4, CSF-1, CCL3, MCP-1, uPA, and CCL19, and consistent downregulation of IL-1α as compared to HC. Notably, mild vs. moderate-and-severe difference reached significance only when severity was defined by total SCORAD (IL-8, CXCL6, CXCL1; all FDR<0.05, (Figure 1C, Table S9)). Likewise, principal component analysis (PCA) of the 24-protein panel (PC1 67.1%, PC2 12.3%) demonstrated separation of HC from both AD groups, with overlap between mild and moderate-and-severe AD across all severity classification schemes (Figure 1D).

**Figure 1.**
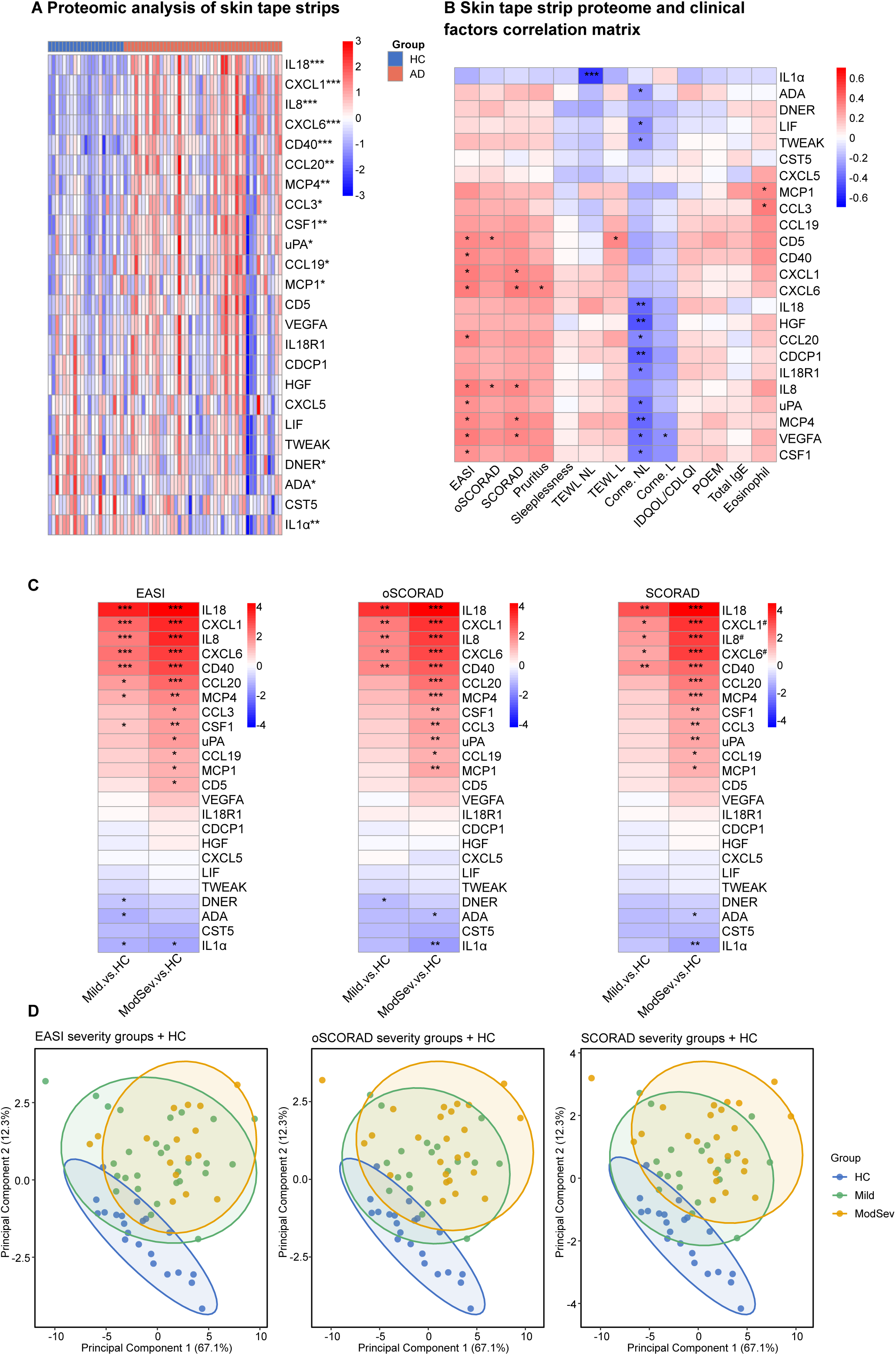
Non-lesional skin in pediatric AD shows an innate-dominated proteomic signature that is present already in mild disease and increases with severity. (A) Heatmap of Z-score normalized NPX values for the 24 detected Olink Inflammation panel proteins in non-lesional STS from HC (n = 21) and patients with AD (n = 44). Proteins are ordered by log₂ fold change (AD vs HC, most upregulated at top). Columns represent individual samples, grouped by study group (HC, blue; AD, red). Asterisks denote FDR-adjusted significance (*limma*, adjusted for age and sex). *FDR < 0.05, **FDR < 0.01, ***FDR < 0.001. (B) Spearman correlation heatmap between STS protein levels and clinical, barrier, and hematological parameters in AD patients only (n = 44). Color indicates the correlation coefficient (ρ); asterisks denote raw (uncorrected) p-values: *p < 0.05, **p < 0.01, ***p < 0.001. (C) Heatmaps of log₂ fold change for mild-versus-HC and moderate and severe (ModSev)-versus-HC comparisons across three severity classification systems, EASI (Mild, n = 28; ModSev, n = 16; HC, n = 21), oSCORAD (Mild, n = 19; ModSev, n = 25; HC, n = 21), and SCORAD (Mild, n = 19; ModSev, n = 25; HC, n = 21). Asterisks denote FDR-adjusted significance (*limma*, adjusted for age and sex): *FDR < 0.05, **FDR < 0.01, ***FDR < 0.001. # - statistically significant difference between Mild and ModSev AD; (D) PCA of STS proteomic profiles in HC (blue, n = 21), Mild AD (green), and moderate and severe AD (orange), shown separately for each severity classification system: EASI (Mild, n = 28; ModSev, n = 16), oSCORAD (Mild, n = 19; ModSev, n = 25), and SCORAD (Mild, n = 19; ModSev, n = 25). Ellipses represent 95% confidence regions. PC1 and PC2 explained 67.1% and 12.3% of total variance, respectively. AD, atopic dermatitis; EASI, Eczema Area and Severity Index; FDR, false discovery rate; HC, healthy controls; PCA, principal component analysis; oSCORAD, objective SCORAD; SCORAD, total SCORAD; TEWL, transepidermal water loss; NL, non-lesional; L, lesional; IDQOL/CDLQI, Infants’ Dermatitis Quality of Life Index/Children’s Dermatology Life Quality Index; POEM, Patient-Oriented Eczema Measure; STS, skin tape strip

### 3.3 Proteomic signatures of non-lesional skin associate with sensitization to foods and co-sensitization

We further stratified AD patients by sensitization pattern: no sensitization (n=5), aeroallergen only (n=9), food only (n=5), and combined food and aeroallergen (n=25), and each was compared with healthy controls (n=21), adjusted for age, sex, and EASI. Neither the non-sensitized nor the aeroallergen-only group differed from controls after FDR correction. In contrast, the food-sensitized group showed significant upregulation of IL-8, IL-18, CXCL1, CXCL6, and CD40 (log_2_FC 2.56 to 3.50), and the co-sensitized (food and aeroallergen sensitized) group showed a comparable signature with additional downregulation of IL-1α and ADA (log_2_FC −1.49 and −1.59) (Figure 2), suggesting that sensitization to foods or co-sensitization can be an additional factor implicated in inflammatory profile of non-lesional skin. No protein differed between any two AD sensitization groups.

**Figure 2.**
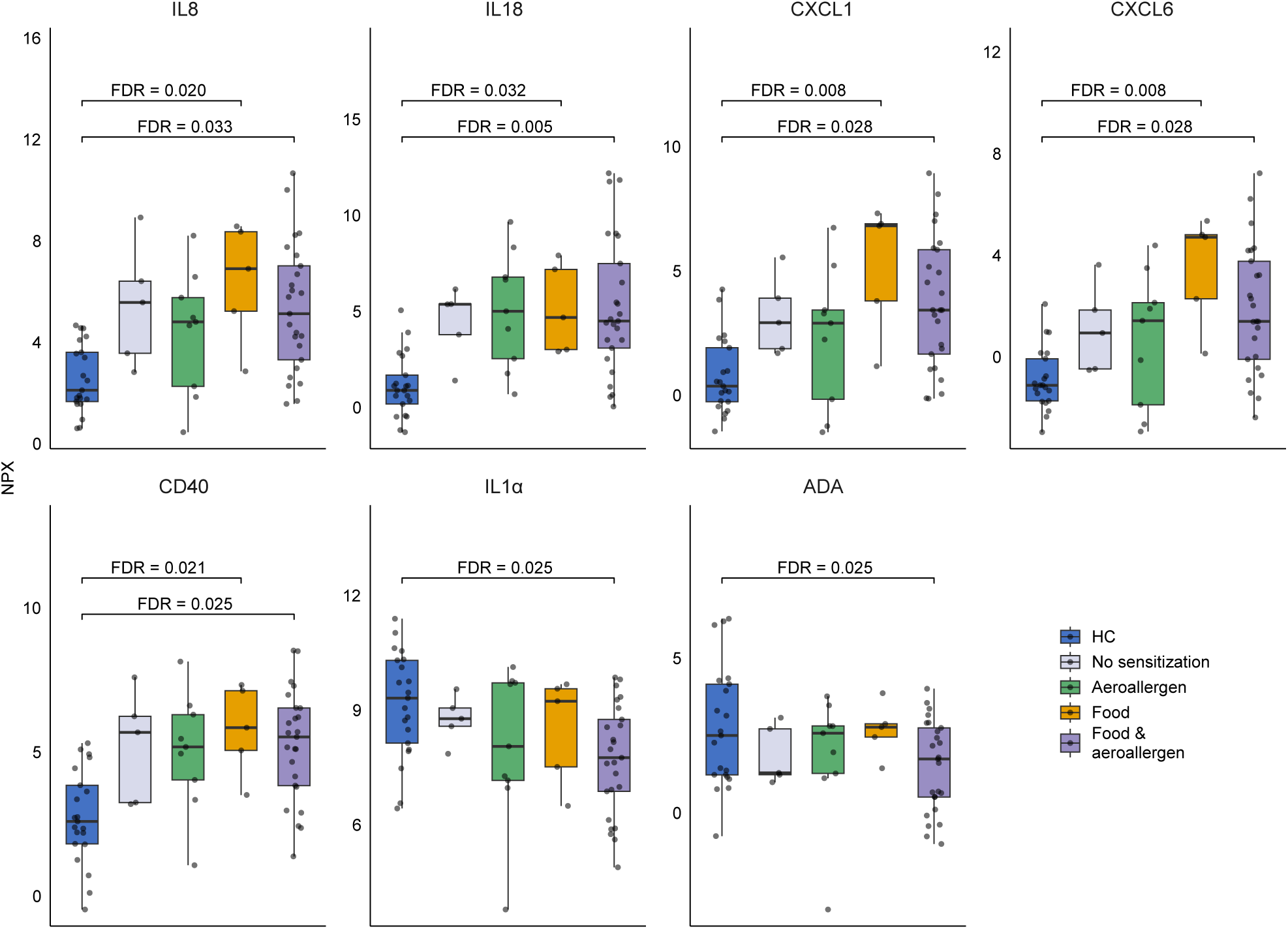
Food- and co-sensitized children differ from healthy controls but not from non-sensitized AD patients in the non-lesional skin tape strip proteome. Boxplots of NPX values for the seven proteins significantly differentially expressed (FDR < 0.05 in at least one comparison) across sensitization groups: HC (n = 21), no sensitization (n = 5), aeroallergen sensitization only (n = 9), food sensitization only (n = 5), and combined food and aeroallergen sensitization (n = 25). Comparisons were performed using the *limma* package in R, adjusted for age, sex, and EASI. Brackets indicate significant comparisons against HC, with exact FDR-adjusted p-values shown. Boxes represent the IQR with the median indicated by the horizontal line; whiskers extend to the most extreme value within 1.5× IQR of the box; individual data points are overlaid. FDR, false discovery rate; HC, healthy controls; IQR, interquartile range

At the level of individual allergens (sensitized and non-sensitized groups compared separately with controls; analysis adjusted for age, sex, and EASI), food allergen sensitization was accompanied by upregulation of the core inflammatory signature, most consistently for storage proteins, tree nuts, and egg compared to HC, each showing significant elevation of IL-8, IL-18, CXCL1, CXCL6, and CD40 (log_2_FC up to 3.33, FDR < 0.05), with visible trends of highest expression in sensitized individuals. However, several proteins were also elevated in the corresponding non-sensitized comparisons against controls, and no protein reached significance in any sensitized-versus-non-sensitized contrast (Figure 3).

**Figure 3.**
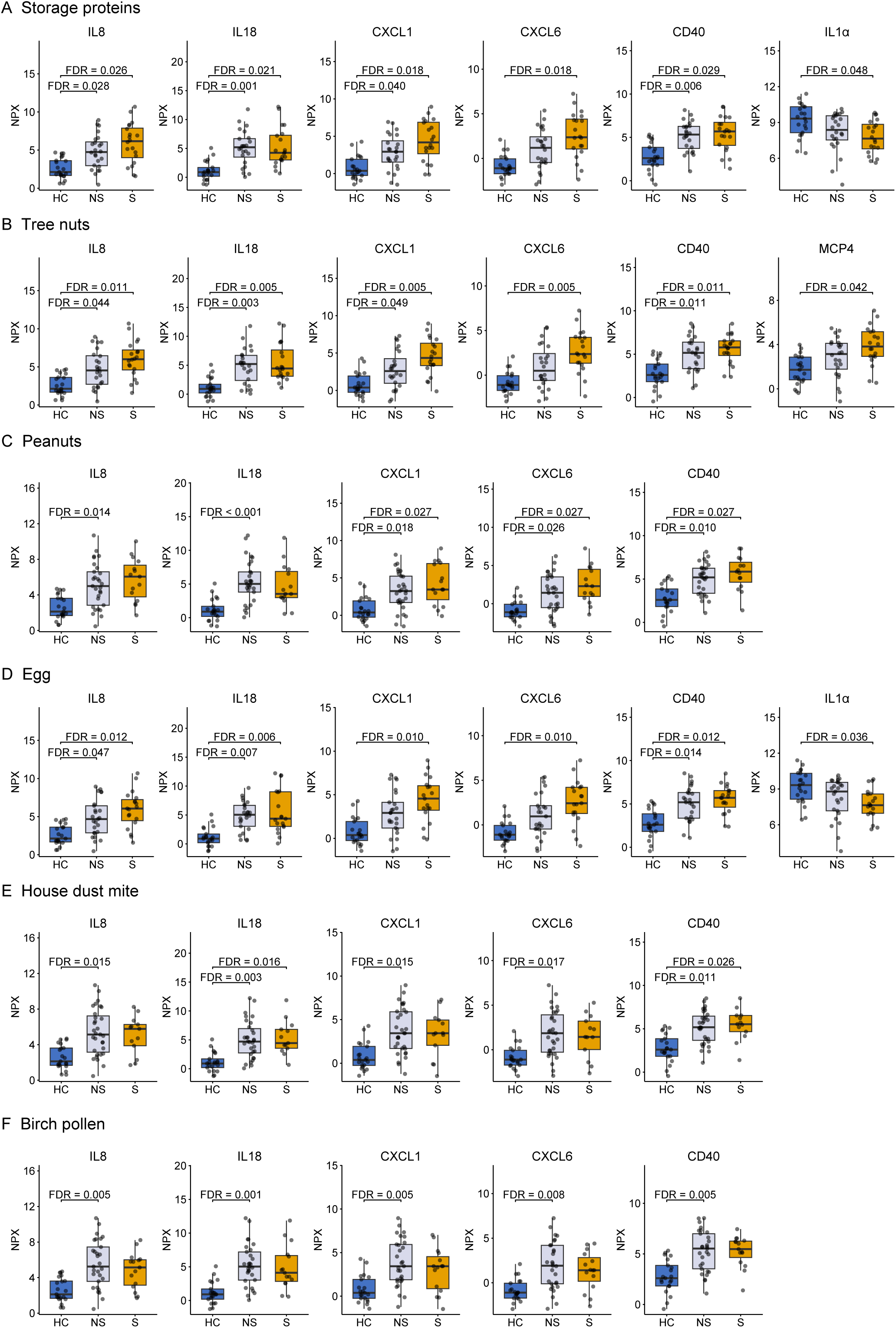
AD patients show elevated innate inflammatory proteins compared with healthy controls, with no differences between sensitized and non-sensitized patients for any allergen. Boxplots of NPX values for proteins significantly differentially expressed for six representative allergen sensitizations: (A) storage proteins (HC, n = 21; non-sensitized, n = 24; sensitized, n = 20); (B) tree nuts (HC, n = 21; non-sensitized, n = 24; sensitized, n = 20); (C) peanuts (HC, n = 21; non-sensitized, n = 29; sensitized, n = 15); (D) egg (HC, n = 21; non-sensitized, n = 25; sensitized, n = 19); (E) house dust mite (HC, n = 21; non-sensitized, n = 31; sensitized, n = 13); (F) birch pollen (HC, n = 21; non-sensitized, n = 29; sensitized, n = 15). For each allergen, patients were classified as non-sensitized or sensitized based on ALEX² component-resolved diagnostics. Comparisons were performed by *limma* package in R, adjusted for age, sex, and EASI. Brackets indicate significant comparisons against HC, with exact FDR-adjusted p-values shown. Boxes represent the IQR with the median indicated by the horizontal line; whiskers extend to 1.5× IQR; individual data points are overlaid. FDR, false discovery rate; HC, healthy controls; IQR, interquartile range; NS, non-sensitized; S, sensitized

### 3.4 Serum proteomic signatures associate with disease severity and egg sensitization

As a next step, we analyzed serum immune markers in children with AD. In the comparison of moderate-and-severe AD versus mild disease (EASI-based, adjusted for age and sex), no protein reached significance after FDR correction in serum. Nevertheless, four proteins showed higher expression in moderate-and-severe disease in non-adjusted p-value: IL-13 (log₂FC = 0.68, p = 0.002, FDR = 0.189), MCP-4 (log₂FC = 0.44, p = 0.013, FDR = 0.497), MCP-3 (log₂FC = 0.30, p = 0.035, FDR = 0.603), and CCL19 (log₂FC = 0.31, p = 0.040, FDR = 0.603) (Figure 4A, Table S10).

**Figure 4.**
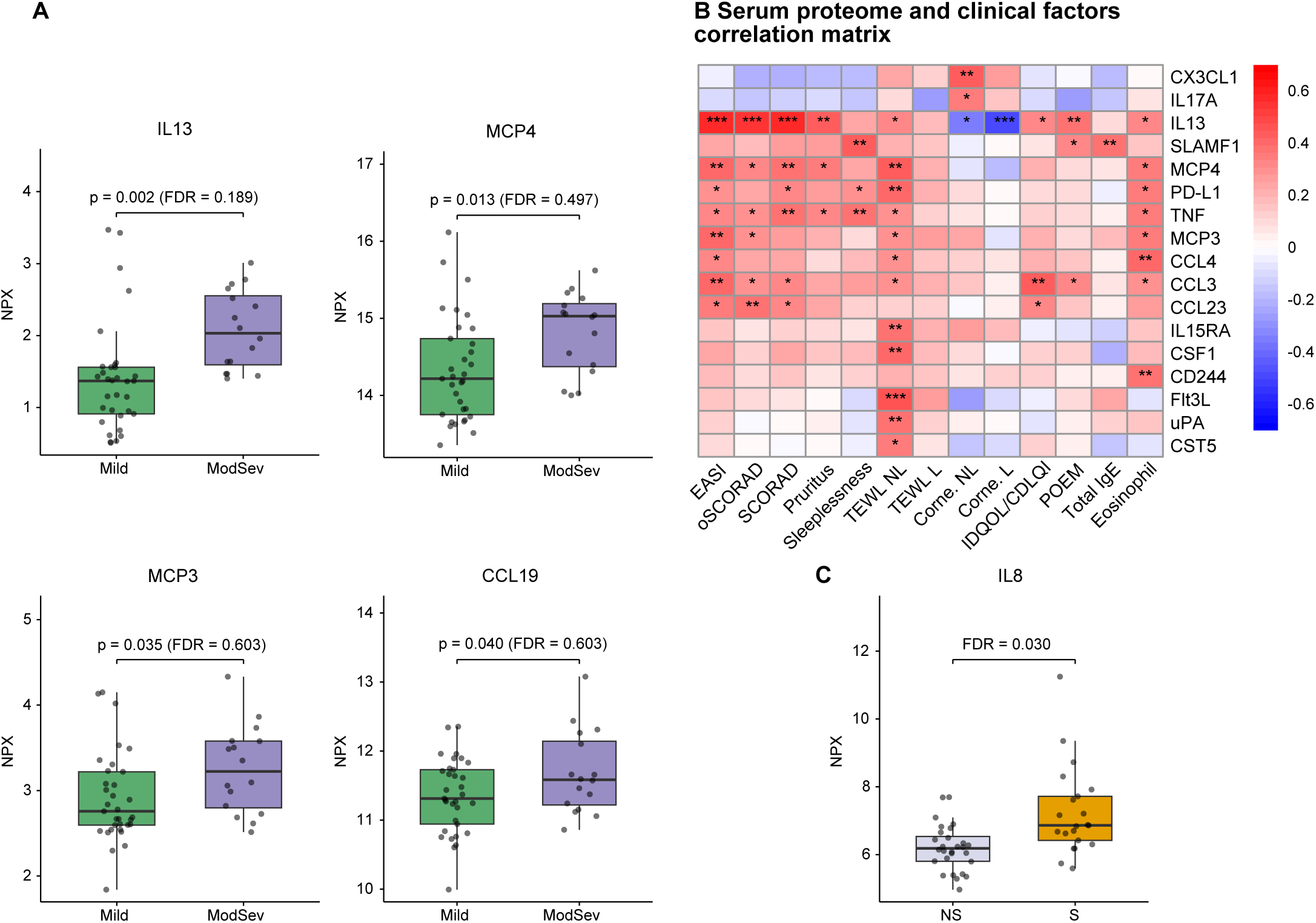
Serum severity associations in pediatric AD are type-2-driven, while egg sensitization is marked by elevated IL-8. (A) Boxplots of NPX values for four proteins with the lowest nominal p-values IL-13, MCP-4, MCP-3, and CCL19 comparing mild (n = 33) and moderate and severe (Mod-Sev, n = 16) AD patients, classified by EASI. Comparisons were performed by *limma* package in R, adjusted for age and sex. Exact p-values and FDR-adjusted p-values are shown. (B) Spearman correlation heatmap between serum protein levels (proteins with |ρ| ≥ 0.35 with at least one clinical variable) and clinical, barrier, and hematological parameters in AD patients (n = 49), unadjusted results. Color indicates the correlation coefficient (ρ); asterisks denote raw (uncorrected) p-values: *p < 0.05, **p < 0.01, ***p < 0.001. (C) Boxplot of NPX values for IL-8 comparing egg non-sensitized (n = 28) and egg-sensitized (n = 21) AD patients. Comparison was performed using the *limma* package in R, adjusted for age, sex, and EASI; the FDR-adjusted p-value is shown. In all panels, boxes represent the IQR with the median indicated by the horizontal line; whiskers extend to 1.5× IQR; individual data points are overlaid. AD, atopic dermatitis; EASI, Eczema Area and Severity Index; FDR, false discovery rate; IQR, interquartile range; NS, non-sensitized; oSCORAD, objective SCORAD; SCORAD, total SCORAD; S, sensitized; TEWL, transepidermal water loss; NL, non-lesional; L, lesional; IDQOL/CDLQI, Infants’ Dermatitis Quality of Life Index/Children’s Dermatology Life Quality Index; POEM, Patient-Oriented Eczema Measure.

In Spearman correlation analyses (unadjusted), IL-13 was the strongest correlate of disease severity, correlating positively with EASI (ρ = 0.59), oSCORAD (ρ = 0.55), and SCORAD (ρ = 0.57; all p < 0.001), pruritus (ρ = 0.43, p = 0.002), and POEM (ρ = 0.38, p = 0.007), and inversely with lesional (ρ = −0.49, p < 0.001) and non-lesional (ρ = −0.35, p = 0.013) corneometry. MCP-4 correlated with EASI (ρ = 0.40; p<0.01), non-lesional transepidermal water loss (ρ = 0.42, p = 0.002), and blood eosinophil count (ρ = 0.36, p = 0.011). Interestingly TEWL in non-lesional skin correlated positively with a broader cytokine signature than in lesional skin (Figure 4B).

Across all sensitization categories, the only serum protein differing significantly after FDR correction was IL-8, which was elevated in egg-sensitized compared with non-sensitized patients (log₂FC = 1.24, p = 0.0004, FDR = 0.03), adjusted for age, sex, and EASI (Figure 4C).

### 3.5. Unsupervised proteomic analysis of non-lesional skin and serum proteome reveals clusters of AD patients

Unsupervised k-means clustering of non-lesional skin targeted proteomic profiles identified two clusters (STS Cluster 1, n = 14; STS Cluster 2, n = 30), separating primarily along the first principal component (PC1 = 72.2% of variance; PC2 = 6.5%) (Figure 5A). Clinically, STS Cluster 2 showed a consistent trend toward greater severity, though without statistical significance (Figure 5E). IL-22 was also higher in STS Cluster 2, with borderline significance (52.29 vs. 24.18 pg/mL, p = 0.064). No other clinical, barrier, or sensitization variable differed between clusters (Table S4). The strongest contributors to PC1 were uPA, VEGFA, CDCP1, IL-18R1, TWEAK, CSF-1, IL-8, and HGF, which were all higher in Cluster 2 (Figure 5C). Bootstrap resampling (B = 1000) confirmed high stability for both STS clusters (Cluster 1 mean Jaccard = 0.915; Cluster 2 mean Jaccard = 0.952). Serum protein expression did not differ between STS-defined clusters for any of the 79 proteins tested (all FDR > 0.97), indicating no detectable systemic proteomic correlate of the local skin inflammatory clustering (Table S11).

**Figure 5.**
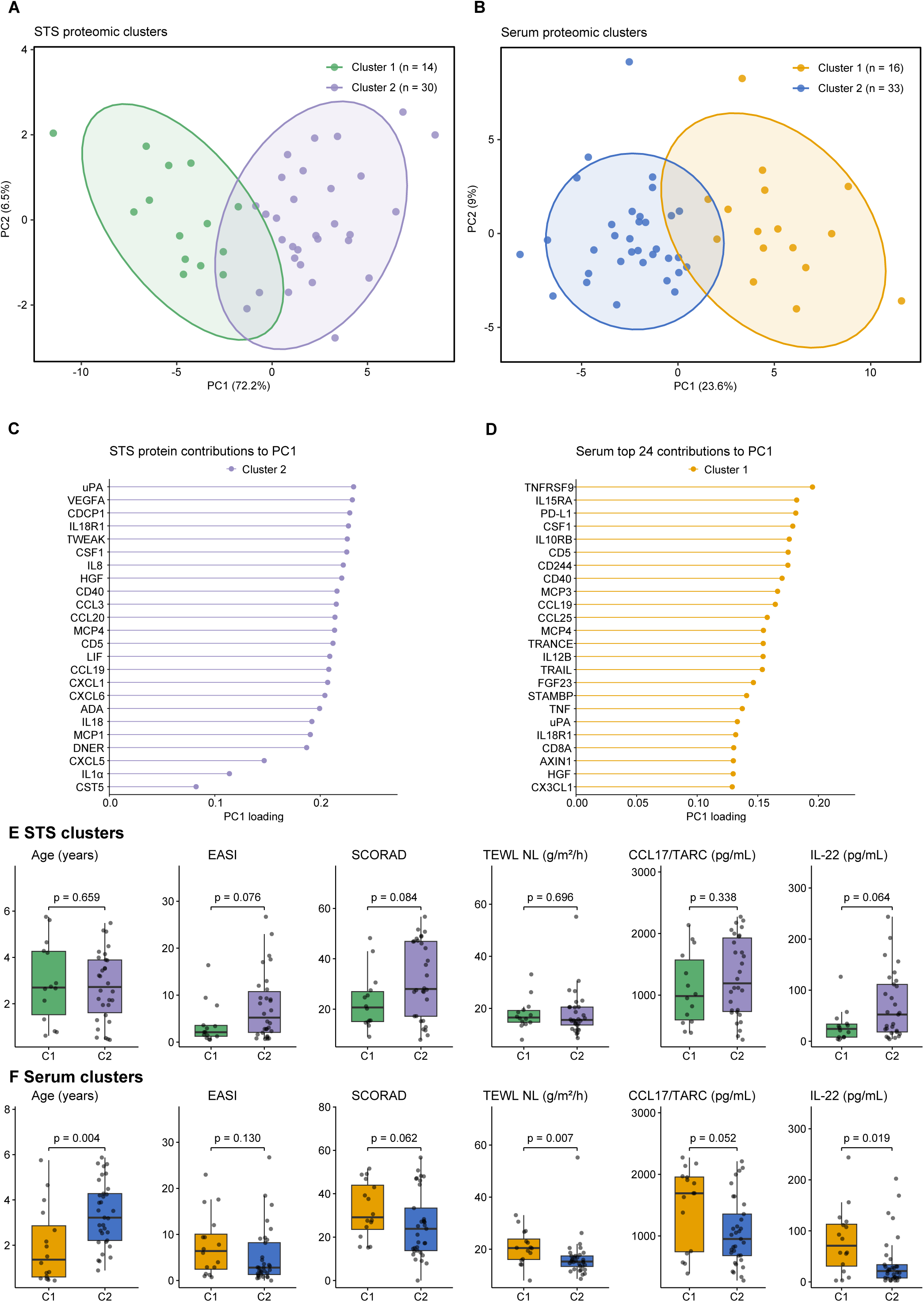
Exploratory proteomic clusters in skin and serum align with different clinical characteristics. (A) Principal component analysis (PCA) of skin tape strip (STS) Olink proteomic profiles in AD patients only (n = 44), colored by k-means cluster assignment (k = 2): Cluster 1 (green, n = 14) and Cluster 2 (purple, n = 30). PC1 and PC2 explained 72.2% and 6.5% of total variance, respectively. (B) Principal component analysis (PCA) of serum Olink proteomic profiles in AD patients only (n = 49), colored by k-means cluster assignment (k = 2): Cluster 1 (orange, n = 16) and Cluster 2 (blue, n = 33). PC1 and PC2 explained 23.6% and 9% of total variance, respectively. For both compartments, ellipses represent 95% confidence regions. (C) Proteins contributing to the separation along PC1 in skin tape strips, ranked by absolute PC1 loading; all 24 detected proteins are shown. (D) The 24 proteins with the highest absolute PC1 loadings in serum, of 79 analyzed. In both panels, color indicates the cluster in which the protein was higher; loadings in the same direction indicate that PC1 reflects the overall magnitude of protein expression rather than a contrast between distinct groups of proteins. (E, F) Clinical and immunological characteristics of the proteomic clusters in skin tape strips (E) and serum (F). Boxes show the median and interquartile range, whiskers extend to 1.5 × IQR, and individual patients are plotted as points. p-values are from the Mann–Whitney U test. C1, Cluster 1; C2, Cluster 2; TEWL NL, transepidermal water loss from non-lesional skin.

Clustering of serum targeted proteomic profiles identified two clusters (Serum Cluster 1, n = 16; Serum Cluster 2, n = 33), separating along the first principal component (PC1 = 23.6% of variance; PC2 = 9%), with small partial overlap between clusters (Figure 5B). Clinically, Serum Cluster 1 was significantly younger (1.36 vs 3.22 years, p = 0.004) and showed greater non-lesional transepidermal water loss (20.47 vs 15.29 g/m²/h, p = 0.007) and higher serum IL-22 (71.00 vs 21.38 pg/mL, p = 0.019) (Figure 5F). Serum Cluster 1 was also strongly enriched for food and aeroallergen sensitization in 87.5% versus 36.4% of patients (p < 0.001), and higher rates of egg sensitization (68.8% vs 30.3%, p = 0.015, Table S4). The strongest contributors to PC1 were TNFRSF9, IL-15RA, PD-L1, CSF-1, IL-10RB, CD5, CD244, and CD40, which were higher in Cluster 1 (Figure 5D). Bootstrap resampling (B = 1000) confirmed high stability for both serum clusters (Cluster 1 mean Jaccard = 0.939; Cluster 2 mean Jaccard = 0.970).

## 4. Discussion

In this study of young children with AD, under 6 years of age, non-lesional skin and serum proteomes showed distinct, compartment-specific immune proteomic clusters. Non-lesional skin showed a robust, severity-graded innate signature that was already present in mild disease, while serum was more heterogeneous, but dominated by a Th2-associated signature that associated with severity. Sensitization to foods and co-sensitization further amplified the innate immune non-lesional skin signature and serum IL-8 was independently associated with egg sensitization. This suggests a modifying, rather than defining, role for sensitization in the immune landscape of early AD.

Consistent with prior tape strip studies in pediatric AD, we found that the non-lesional skin proteome was substantially altered relative to healthy skin, with the strongest signal carried by innate immune proteins (6,8,12). IL-8, IL-18, CXCL1, CXCL6, CD40 (upregulated), and IL-1α (downregulated) were among the proteins that most consistently differentiated children with AD from HC, and several also tracked disease severity (13). This pattern is in line with previous studies identifying innate immune mediators in non-lesional skin as biomarkers of barrier dysfunction and AD severity (6,13,14), yet the fact that this signature can be observed already in mild disease and is further amplified in moderate-and-severe disease extends prior work.

Among significantly upregulated cytokines, IL-18 warrants particular attention. IL-18, an IL-1-family alarmin, stored as pro-IL-18 in keratinocytes and released upon epithelial injury or inflammasome activation (15) is increasingly recognized as a driver of type 2 skewing in the skin, amplifying allergic inflammation rather than opposing it (16). Elevated skin and serum IL-18 correlate with AD severity across multiple cohorts (17–19), and IL-18 measured in clinically normal skin of neonates has been reported to predict later moderate and severe AD, supporting its candidacy as an early and severity-tracking biomarker (20).

While innate immune mediators dominated the skin severity signal, the serum severity was nominally associated with Th2-related markers - IL-13, the canonical Th2 effector cytokine and the Th2-associated chemokines MCP-3 and MCP-4, as previously described in similar cohorts (21,22). Notably, MCP-4 showed a nominal trend toward higher levels in serum of moderate-and-severe AD and was significantly elevated in non-lesional skin of patients with moderate-and-severe AD, suggesting that a Th2-associated signal is not confined to the systemic compartment but is also detectable locally.

We next evaluated whether predefined sensitization patterns or sensitization to specific allergens were associated with the non-lesional skin proteomic signature. Children co-sensitized to food and aeroallergens, and those sensitized to food only, showed the same core signature as the AD group as a whole relative to HC, but did not differ from other AD sensitization groups. This is partly consistent with the MPAACH cohort, in which co-sensitization was linked to barrier dysfunction (23), although not to keratinocyte inflammatory markers (24).

At the level of individual allergens, CXCL6 was selectively increased in patients sensitized to tree nuts, egg, and storage proteins and IL-1α selectively decreased in patients sensitized to egg and storage proteins, compared with HC. In patients sensitized to the aforementioned allergens, only a trend toward increased IL-8, CXCL1, CXCL6 and CD40 was observed compared to non-sensitized AD patients. Leung, in contrast, described a distinct non-lesional skin proteomic endotype in peanut-allergic AD-children relative to non-allergic AD, correlating with polysensitization in the same cohort (25,26). In our cohort, no protein clearly distinguished sensitized from non-sensitized patients. A subset of proteins did, however, distinguish specific allergen-sensitized groups from healthy controls. An allergen-specific signal may therefore be emerging within a broadly shared inflammatory background, though confirmation will require future investigation. Furthermore, children sensitized to egg showed higher IL-8 levels in serum, when adjusted for age, sex and AD severity. Sensitization to food allergens is classically understood to proceed via a dendritic cell (DC)–driven Th2 pathway, culminating in IgE-mediated sensitization of mast cells and basophils (27–29). On the other hand, IL-8 has been linked to allergic activation in food allergy through IgE-mediated mast cell degranulation (30), and CXCL8 transcript levels in PBMCs of egg-allergic children have been reported to decline during oral immunotherapy (31), providing some precedent for an IL-8-related innate contribution to food allergy. Taken together, our findings suggest that the dominant proteomic signal is driven by AD itself. Additional contribution of allergen sensitization appears limited to serum IL-8 differential expression in egg sensitized individuals. Sensitization-related findings relative to healthy controls should be regarded as exploratory and require independent validation due to small sample size.

Finally, unsupervised clustering performed separately for STS and serum was used to explore whether proteomic subgroups exist independently of pre-specified clinical categories. In skin, cluster assignment was, by construction, driven by the same core inflammatory proteins identified in the severity analyses, and the resulting clusters showed a corresponding trend toward higher severity in the more inflamed cluster (STS Cluster 2), without differing in sensitization profile. The STS clusters differed in sex distribution at borderline significance (Table S4), a finding we cannot explain and that warrants caution.

In serum, two clusters emerged with distinct clinical and immunological profiles, reflecting a partially age- and severity-related immune pattern: a younger, barrier-impaired, more sensitized, Th22-skewed phenotype (Serum Cluster 1) versus an older, less inflamed, aeroallergen-predominant phenotype (Serum Cluster 2), rather than a clean division along disease severity alone. This raises the possibility that Serum Cluster 1 reflects an age-associated immunological stage of disease and association with co-sensitization. Bakker et al. identified four biomarker-based clusters in pediatric AD, of which one was severity-dependent while the remaining clusters were defined by specific immune signatures (32). In our cohort, by contrast, age and sensitization burden discriminated better than severity, although comparison is limited by our much smaller cohort and narrower age range (<6 vs. up to 17 years).

Cross-compartment analysis found no significant difference in serum protein expression between STS-defined clusters. Given the limited sample size, this null result does not establish that the two compartments are independent, but it is consistent with the broader observed pattern in which severity in skin was associated with innate mediators and in serum with type-2 markers, and the two cluster solutions aligned with different clinical variables. Taken together, these observations support a model in which non-lesional skin and serum proteomes capture distinct, only partially overlapping dimensions of pediatric AD heterogeneity.

This study profiles paired skin and serum proteomes across the pediatric AD severity spectrum, including mild disease, enabling direct comparison between compartments within the same children. Sampling non-lesional skin revealed molecular changes beyond visible lesions, while ALEX² provided a component-resolved definition of sensitization. The smaller subgroups and absence of external validation mean that the identified profiles warrant confirmation in larger cohorts. Their stability and causal relationships also require longitudinal study. ALEX² sensitization was not linked to confirmed clinical allergy, and controls were not screened for subclinical sensitization. Although only 24 proteins were reliably detected in skin tape strips, limiting assessment of low-abundance type 2 mediators, the paired samples provide a valuable basis for investigating skin and systemic immune patterns in early childhood AD.

Immune profiles in early pediatric AD appear to be primarily shaped by disease severity, with a limited contribution from allergen sensitization. Skin and serum thus appear to be complementary rather than interchangeable sources of biomarker information in this age group. Confirming whether the profiles identified in this narrow developmental window represent stable biological subgroups will require larger, longitudinally followed cohorts with external validation.

## Supporting information

Supplementary materials part 1

Supplementary materials part 2

## Online Repository

Yes

## Abbreviations

AD: Atopic dermatitis
ALEX²: Allergy Explorer
CDLQI: Children’s Dermatology Life Quality Index
DC: Dendritic cell
EASI: Eczema Area and Severity Index
FcεRI: High-affinity IgE receptor
FDR: False discovery rate
HC: Healthy control(s)
HDM: House dust mite
IDQOL: Infants’ Dermatitis Quality of Life index
NL: Non-lesional
NPX: Normalized protein expression
NS: Non-sensitized
oSCORAD: Objective SCORAD
PCA: Principal component analysis
POEM: Patient-Oriented Eczema Measure
S: Sensitized
SCORAD: SCORing Atopic Dermatitis (total SCORAD)
STS: Skin tape strip(s)
TEWL: Transepidermal water loss
VAS: Visual Analog Scale

## Data Availability

The data that support the findings of this study are available from K.D. or W.F. upon reasonable request.

