## Supplementary materials part 1 for "Paired skin and serum proteomics capture distinct immune patterns in early childhood atopic dermatitis"

**Short title:** Skin and Serum Proteomics in Pediatric AD

**Authors:**

Karolina Dumycz^1,2,3^, Philipp Gessner^3,4,5^, Joanna Nowiszewska^1,2,6^, Dominika Ambrożej^1,2^, Anja Heider^3^, Anna Stelmaszczyk-Emmel^7^, Dominika Nowis^8^, Cezmi Akdis^3^, Marek Kulus^1^, Christoph Messner^3,4,5,9^, Milena Sokolowska^3*^, Wojciech Feleszko^1*^

1. Department of Pediatric Pneumonology and Allergy, Medical University of Warsaw, Warsaw, Poland

2. Doctoral School, Medical University of Warsaw, Warsaw, Poland

3. Swiss Institute of Allergy and Asthma Research (SIAF), University of Zurich, Davos, Switzerland

4. Precision Proteomics Center, University of Zurich, 7265 Davos, Switzerland

5. Swiss Institute of Bioinformatics (SIB), 1005 Lausanne, Switzerland

6. Department of Dermatology, Medical University of Warsaw, Warsaw, Poland

7. Department of Laboratory Diagnostics and Clinical Immunology of Developmental Age, Medical University of Warsaw

8. Laboratory of Experimental Medicine, Medical University of Warsaw, Warsaw, Poland

9. The LOOP Zurich, 8044 Zurich

* These authors should be considered as shared senior authors

**METHODS**

**Clinical assessment**

Quality of life was assessed using two age-appropriate instruments: the Infants' Dermatitis Quality of Life Index (IDQOL) in children under 4 years of age and the Children's Dermatology Life Quality Index (CDLQI) in children aged 4 years and above. Both instruments comprise 10 items scored on an identical 0–3 scale, yielding a total score ranging from 0 to 30, with higher scores indicating greater impairment of quality of life. Because the two questionnaires share the same structure, scoring range, and interpretive framework, and differ only in the age-adapted wording of individual items, scores were combined into a single variable (IDQOL/CDLQI) for analysis, with the instrument applied determined by the child's age at assessment.

**Skin barrier assessment**

Skin barrier function was evaluated by measuring transepidermal water loss (TEWL) and stratum corneum hydration. TEWL was assessed using the Tewameter® TM 300 (Courage + Khazaka Electronic GmbH, Cologne, Germany) on lesional and non-lesional skin of the upper extremities, preferably on the antecubital fossa and forearm in standardized humidity and temperature of the examination room (21.9-23.5°C; 40-65% humidity). Stratum corneum hydration was measured using the Corneometer® CM 825 (Courage + Khazaka Electronic GmbH) on the same skin regions. A minimum of three to four readings were performed per site for each measurement type, and the average value was used in analysis.

**Skin tape-stripping**

Minimally invasive epidermal sampling was performed using the skin tape-stripping (STS) technique. Up to fifteen consecutive D-Squame® Sampling Discs (D102, Clinical & Derm LLC) were applied to non-lesional skin preferably on the volar forearm using a standardized D-Squame® Pressure Instrument (D500, Clinical & Derm LLC) to ensure uniform pressure. After each application, tape-strips were removed using gloved hands and tweezers and placed individually onto D-Squame® Standard Storage Cards (D120, Clinical & Derm LLC). Samples were immediately stored at −80°C until further analysis.

**Serum collection**

Peripheral blood samples (approximately 2 mL) were collected into tubes containing clot activator. After allowing the blood to clot at room temperature for 30–120 minutes, samples were centrifuged at 2,000 × g for 10 minutes. The resulting serum was aliquoted into at least four cryotubes (minimum 150 μL per tube) and stored at −80°C for subsequent analyses.

**Protein Extraction from Skin Tape-Strips**

Protein extraction from stratum corneum was performed using the tape-strips collected as described above. For each participant, four tape-strips with matching numbers (strips no. 3, 6, 7, and 8) were used for protein extraction. Extraction was carried out using radioimmunoprecipitation assay (RIPA) buffer supplemented with protease inhibitors. Briefly, 1X RIPA buffer was prepared from a 10X stock solution (Millipore, #20-188) using distilled water and supplemented with a protease inhibitor cocktail (1 tablet of cOmplete™ Mini, Roche) per 10 mL of buffer. Tape-strips were thawed on ice and individually placed into 1.5 mL sterile microcentrifuge tubes using sterile tweezers, ensuring each strip was fully positioned at the bottom of the tube and did not interfere with the tube cap seal. A total of 600 µL of prepared 1× RIPA buffer was added to the first tape-strip (strip no. 3), and the tube was subjected to 15 minutes of sonication in an ice-cooled ultrasonic water bath. After brief centrifugation (15,000 × g, 4°C, 1 minute), the buffer was carefully transferred to a second tube containing the next strip (no. 6), and the sonication process was repeated. This serial extraction was continued across all four strips to maximize protein yield. Following the final sonication, the pooled extract was transferred to a fresh tube and centrifuged at 15,000 × g for 5 minutes at 4°C to remove particulate debris. The supernatant was aliquoted and stored at −80°C until analysis. A 60 µL aliquot was reserved for protein quantification. Total protein concentration was determined using the BCA Protein Assay Kit (Pierce™ BCA Protein Assay, Thermo Fisher Scientific) according to the manufacturer’s instructions.

**Proteomic analysis of skin tape strips and serum**

Quantitative proteomic profiling was performed on both serum samples and protein extracts obtained from skin tape-strips using the Olink® Target 96 Inflammation panel (Olink Proteomics, Uppsala, Sweden). This proximity extension assay (PEA)-based technology enables high-throughput, multiplexed detection of 92 inflammation-related protein biomarkers with high specificity and sensitivity. A 10 µL of serum and 10 µL of skin protein extract per individual were submitted for analysis. All samples were randomized, and assays were performed according to the manufacturer’s standardized protocol at a certified Olink service facility. Protein expression levels were reported in Normalized Protein eXpression (NPX) units on a log2 scale, the standard normalization method provided by Olink for its proximity extension assay platform, allowing for relative quantification and comparison across samples within the same batch. Internal controls provided by Olink were used to ensure analytical precision and data quality. All STS samples and all serum samples were analyzed in one batch per material, precluding between-batch variation within each material. Proteins detected in less than 60% of the samples were excluded from further statistical analysis to minimize the impact of missing data and enhance the robustness of the findings. For individuals with missing data, imputation was carried out using the *“missForest”* package, which applies a non-parametric random forest-based method suitable for mixed-type data. Additionally, serum TARC/CCL17 and IL-22 were measured using Human Quantikine ELISA kits (R&D systems) according to manufacturer’s instructions. Serum samples below detection rate were imputed with the half of the value of lower detection limit.

**Statistical Analysis**

All statistical analyses were performed using R software (version 4.3.3). Protein expression data obtained from skin tape-strip samples were normalized to the total protein concentration as determined by the BCA assay to account for inter-sample variability in protein yield according to Olink instructions. Proteins detected in fewer than 60% of all samples were excluded from the dataset prior to analysis. **Patients were classified as sensitized to a given allergen if they demonstrated IgE reactivity to at least one molecular component or the corresponding allergen extract included in the ALEX^2^ panel. Two distinct clustering approaches were applied. First, PCA of skin tape-strip proteomic data was performed using *cluster* package and patients were subsequently classified according to pre-defined severity groups (HC, mild, moderate and severe).** Second, unsupervised k-means clustering was performed independently on STS and serum Olink proteomic data to identify exploratory proteomic subgroups within the AD cohort. Prior to analysis, NPX values for all proteins were Z-score normalized (centered and scaled to unit variance) across patients to ensure equal protein contribution to Euclidean distance. To reduce dimensionality and mitigate the curse of dimensionality inherent to high-dimensional proteomic data, principal component analysis (PCA) was first applied to the Z-scored matrix. The number of principal components (PCs) retained for clustering was determined by the cumulative variance threshold criterion: the minimum number of PCs required to explain ≥80% of total variance was selected. K-means clustering was applied to the retained PC scores. The optimal number of clusters was determined using three complementary approaches: the elbow method (within-cluster sum of squares), the average silhouette width method (Figure S1, S2), **both implemented using the *factoextra* package** and the gap statistic with 100 bootstrap reference sets, **computed using the *cluster* package.** Among solutions with k ≥ 2, all three criteria favored k = 2; the average silhouette width at k = 2 was 0.50 for STS and 0.21 for serum (Figures S1–S2), indicating reasonable cluster structure in STS but only weak structure in serum. K-means was run with 50 random initializations (nstart = 50) and a fixed random seed (123) to ensure reproducibility. Cluster stability was assessed by bootstrap resampling using the *clusterboot* function from the *fpc* package (B = 1000 bootstrap resamples). Cluster robustness was quantified by the mean Jaccard similarity coefficient between bootstrap cluster assignments and the original partition; values >0.85 indicate highly stable clusters, 0.75–0.85 stable, 0.60–0.75 borderline, and <0.60 unstable. Clusters were visualized by projecting patient coordinates onto the first two principal components (PC1 and PC2) with 95% confidence ellipses. Cluster characterization was performed by comparing clinical and immunological variables between clusters using the Mann-Whitney U test for continuous variables and Fisher's exact test for categorical variables; p-values are presented without correction for multiple comparisons, as these analyses are descriptive and exploratory in nature. Proteins contributing to cluster separation were identified from the PCA loadings on the principal component along which clusters separated (PC1). Loadings were ranked by absolute value, and the direction of each loading was interpreted relative to the mean PC1 score of each cluster. Clustering procedure, including principal component selection, k-means clustering, and bootstrap-based Jaccard stability assessment, was applied independently and identically to the skin tape-strip and serum proteomic datasets.

Differential Olink protein expression analysis was performed using the *limma* package (Linear Models for Microarray Data). Linear models were adjusted for age, sex and in some cases EASI as covariates to control for potential confounding effects. Statistical significance was defined at a false discovery rate (FDR) < 0.05 unless otherwise specified. Differences in continuous clinical parameters between groups were assessed using the Mann–Whitney U test. Associations between categorical variables were evaluated using the chi-square test or Fisher's exact test, as appropriate based on expected cell frequencies. Correlations between continuous variables were assessed using Spearman's rank correlation coefficient. All visualizations and plots were generated using standard RStudio packages.

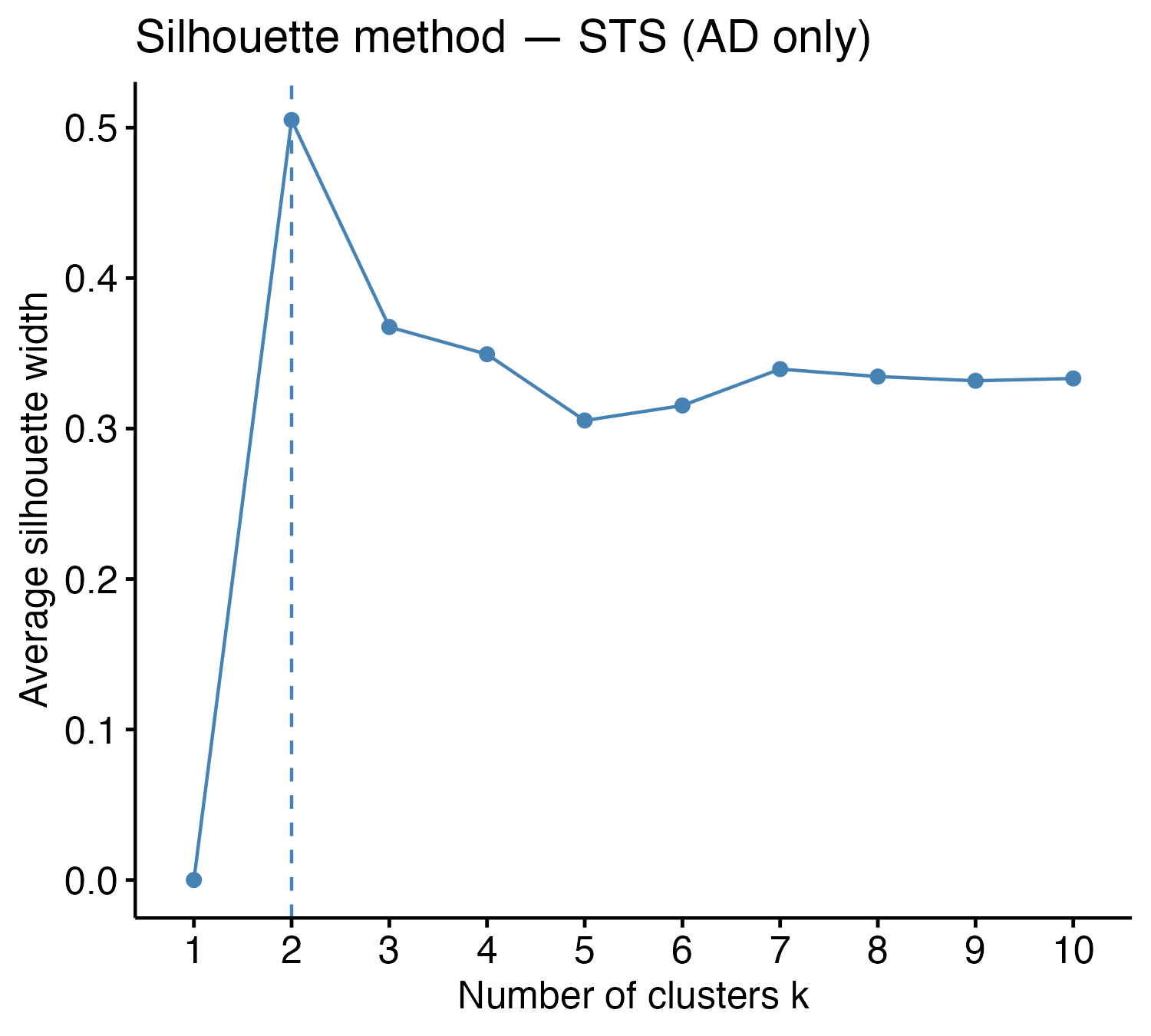

**Figure S1.** Average silhouette width across k = 1–10 for skin tape strip Olink proteomic k-means clustering, AD patients only. Peak silhouette width (0.50) is observed at k = 2.

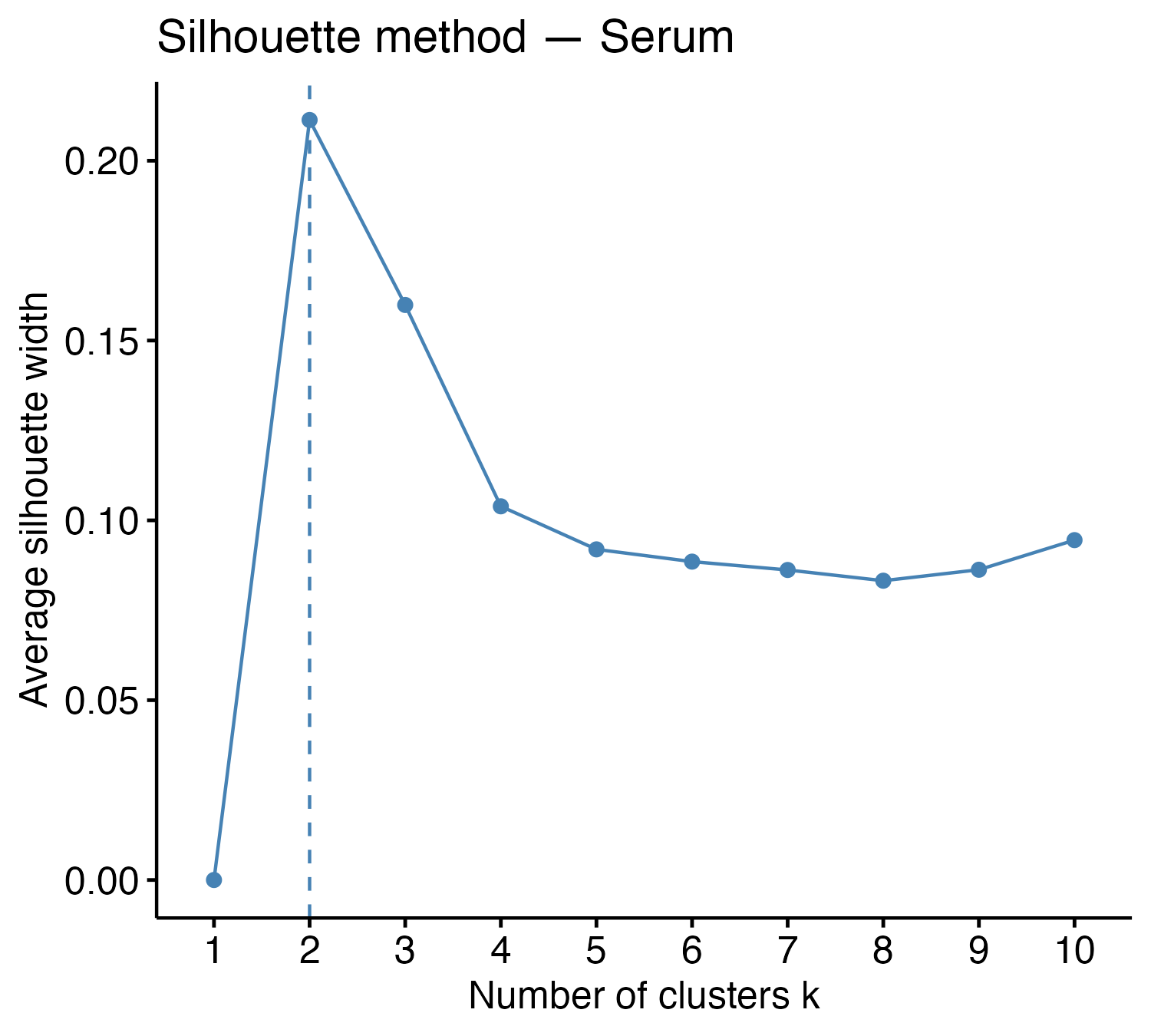

**Figure S2.** Average silhouette width across k = 1–10 for serum Olink proteomic k-means clustering. Peak silhouette width (0.21) is observed at k = 2.

**RESULTS**

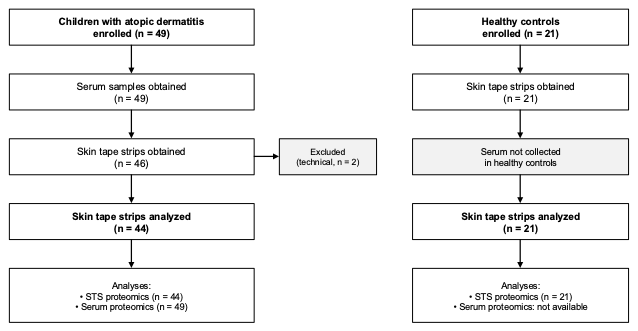

**Figure S3.** Study flow diagram.

**Table S1.** Sensitization profile of AD patients stratified by EASI-based severity group

| **Variable** | **AD Mild (n=33)** | **AD ModSev (n=16)** | **p-value** |
| --- | --- | --- | --- |
| **Type of sensitization, n (%)** |  |  |  |
| Aeroallergen sensitization only | 7 (21.2%) | 3 (18.8%) | 1.000 |
| Food & aeroallergen sensitization | 15 (45.5%) | 11 (68.8%) | 0.143 |
| Food sensitization only | 7 (21.2%) | 1 (6.2%) | 0.245 |
| No sensitization | 4 (12.1%) | 1 (6.2%) | 1.000 |
| **Specific allergen sensitization, n (%)** |  |  |  |
| PR-10 | 11 (33.3%) | 9 (56.2%) | 0.215 |
| Storage proteins (plants) | 14 (42.4%) | 10 (62.5%) | 0.232 |
| Peanuts | 9 (27.3%) | 8 (50.0%) | 0.200 |
| Tree nuts | 11 (33.3%) | 10 (62.5%) | 0.070 |
| Milk | 6 (18.2%) | 4 (25.0%) | 0.709 |
| Egg | 13 (39.4%) | 8 (50.0%) | 0.547 |
| Fish | 3 (9.1%) | 3 (18.8%) | 0.377 |
| Seafood | 5 (15.2%) | 2 (12.5%) | 1.000 |
| Wheat | 2 (6.1%) | 3 (18.8%) | 0.313 |
| Soy | 3 (9.1%) | 6 (37.5%) | 0.043 |
| HDM | 7 (21.2%) | 6 (37.5%) | 0.304 |
| Birch pollen | 8 (24.2%) | 8 (50.0%) | 0.106 |
| Animal fur | 14 (42.4%) | 9 (56.2%) | 0.542 |

Data are presented as n (%). p-values reflect comparison between AD Mild and AD ModSev (Fisher's exact test applied independently for each sensitization category). Sensitization assessed using the ALEX^2^ component-resolved diagnostic platform. HDM: house dust mite

**Table S2.** Hanifin & Rajka minor criteria in AD patients stratified by EASI-based severity group

| **Variable** | **AD Mild (n=33)** | **AD ModSev (n=16)** | **p-value** |
| --- | --- | --- | --- |
| Xerosis | 27 (81.8%) | 16 (100.0%) | 0.159 |
| Palmar hyperlinearity | 8 (24.2%) | 12 (75.0%) | 0.001 |
| Keratosis pilaris | 3 (9.1%) | 3 (18.8%) | 0.377 |
| Hand/foot eczema | 9 (27.3%) | 7 (43.8%) | 0.333 |
| Nipple eczema | 1 (3.0%) | 0 (0.0%) | 1.000 |
| Cheilitis | 0 (0.0%) | 1 (6.2%) | 0.327 |
| Dennie-Morgan fold | 26 (78.8%) | 16 (100.0%) | 0.080 |
| Orbital darkening | 8 (24.2%) | 6 (37.5%) | 0.501 |
| Facial pallor | 12 (36.4%) | 9 (56.2%) | 0.228 |
| Pityriasis alba | 1 (3.0%) | 1 (6.2%) | 1.000 |
| Anterior neck fold | 1 (3.0%) | 2 (12.5%) | 0.245 |
| Perifollicular accentuation | 0 (0.0%) | 1 (6.2%) | 0.327 |
| White dermographism | 13 (39.4%) | 13 (81.2%) | 0.007 |

Data are presented as n (%). p-values reflect comparison between AD Mild and AD ModSev (Fisher's exact test). Minor criteria assessed according to Hanifin & Rajka

**Table S3.** Disease history, environmental factors, and parental atopy stratified by EASI-based severity group

| **Variable** | **AD Mild (n=33)** | **AD ModSev (n=16)** | **p-value** |
| --- | --- | --- | --- |
| **Age of onset, n (%)** |  |  |  |
| 0–3 months | 10 (30.3%) | 9 (56.2%) | 0.119 |
| 3–6 months | 9 (27.3%) | 4 (25.0%) | 1.000 |
| 6–12 months | 7 (21.2%) | 0 (0.0%) | 0.080 |
| >12 months – 2 years | 3 (9.1%) | 2 (12.5%) | 1.000 |
| >2 – 6 years | 4 (12.1%) | 1 (6.2%) | 1.000 |
| Bacterial skin infections (ever) | 10 (30.3%) | 7 (43.8%) | 0.523 |
| Eczema herpeticum (ever) | 0 (0.0%) | 3 (18.8%) | 0.030 |
| Number of initial skin sites | 5.00 (3.00–6.00) | 3.00 (2.00–4.00) | 0.019 |
| Exclusive breastfeeding ≥3 months | 26 (78.8%) | 12 (75.0%) | 1.000 |
| Day care attendance | 18 (54.5%) | 7 (43.8%) | 0.551 |
| Animals at home (currently) | 17 (51.5%) | 6 (37.5%) | 0.382 |
| Cigarette smoke exposure (currently) | 3 (9.1%) | 2 (12.5%) | 1.000 |
| Animals at home (first 2 years) | 18 (54.5%) | 5 (31.2%) | 0.143 |
| Cigarette smoking in pregnancy | 0 (0.0%) | 0 (0.0%) | 1.000 |
| Parental atopy (either parent) | 17 (51.5%) | 10 (62.5%) | 0.549 |
| Paternal atopy | 7 (21.2%) | 4 (25.0%) | 1.000 |
| Maternal atopy | 13 (39.4%) | 7 (43.8%) | 1.000 |

Data are presented as n (%) for categorical variables and median (IQR) for continuous variables. p-values reflect comparison between AD Mild and AD ModSev (Fisher's exact test for categorical variables; Mann-Whitney U test for number of initial skin sites).

**Table S4.** Clinical and immunological characteristics of skin tape strip and serum proteomic clusters

| **Variable** | **Skin tape strip** | | | **Serum** | | |
| --- | --- | --- | --- | --- | --- | --- |
|  | **Cluster 1**  **(n = 14)** | **Cluster 2**  **(n = 30)** | **p-value** | **Cluster 1**  **(n = 16)** | **Cluster 2**  **(n = 33)** | **p-value** |
| Age (years) | 2.70 (1.53-4.26) | 2.73 (1.62-3.89) | 0.659 | 1.36 (0.60-2.86) | 3.22 (2.21-4.28) | **0.004** |
| Sex (female), n (%) | 3 (21.4%) | 17 (56.7%) | 0.050 | 8 (50.0%) | 14 (42.4%) | 0.761 |
| EASI | 2.10 (1.25-3.55) | 5.20 (2.08-10.75) | 0.076 | 6.40 (2.45-10.05) | 2.80 (1.30-8.20) | 0.130 |
| oSCORAD | 14.96 (11.01-17.05) | 21.48 (11.54-33.95) | 0.110 | 19.07 (14.62-33.31) | 15.47 (10.78-23.87) | 0.205 |
| SCORAD | 20.68 (15.10-26.98) | 28.00 (17.21-46.91) | 0.084 | 29.15 (23.56-43.96) | 23.85 (13.78-33.40) | 0.062 |
| IDQOL/CDLQI | 6.00 (5.00-8.00) | 8.50 (4.00-12.75) | 0.471 | 7.00 (5.00-12.50) | 6.00 (4.00-10.00) | 0.244 |
| TEWL NL (g/m²/h) | 16.52 (14.73-19.21) | 15.58 (13.67-20.52) | 0.696 | 20.47 (16.08-23.91) | 15.29 (13.41-17.44) | **0.007** |
| TEWL L (g/m²/h) | 34.64 (26.52-39.44) | 36.04 (30.06-44.67) | 0.385 | 38.58 (34.04-45.16) | 34.00 (28.45-42.99) | 0.321 |
| Eosinophil count (10³/µL) | 0.43 (0.28-0.55) | 0.53 (0.35-0.71) | 0.267 | 0.52 (0.41-0.92) | 0.39 (0.24-0.62) | 0.067 |
| Total IgE (kU/L) | 104.80 (36.88-162.00) | 111.50 (44.38-662.00) | 0.385 | 134.00 (37.85-578.00) | 114.00 (80.90-204.00) | 0.733 |
| CCL17/TARC (pg/mL) | 983.55 (597.28-1570.81) | 1191.22 (732.82-1927.67) | 0.338 | 1692.42 (741.97-1956.13) | 950.17 (679.05-1355.60) | 0.052 |
| IL-22 (pg/mL) | 24.18 (8.02-33.60) | 52.29 (18.27-111.39) | 0.064 | 71.00 (31.02-113.11) | 21.38 (7.89-33.71) | **0.019** |
| **Sensitization profile** | | | | | | |
| No sensitization, n (%) | 2 (14.3%) | 3 (10.0%) | 0.647 | 0 (0.0%) | 5 (15.2%) | 0.158 |
| Food sensitization, n (%) | 1 (7.1%) | 4 (13.3%) | 1.000 | 2 (12.5%) | 6 (18.2%) | 1.000 |
| Aeroallergen sensitization, n (%) | 3 (21.4%) | 6 (20.0%) | 1.000 | 0 (0.0%) | 10 (30.3%) | **0.020** |
| Food and aeroallergen sensitization, n (%) | 8 (57.1%) | 17 (56.7%) | 1.000 | 14 (87.5%) | 12 (36.4%) | **<0.001** |
| Egg, n (%) | 4 (28.6%) | 15 (50.0%) | 0.211 | 11 (68.8%) | 10 (30.3%) | **0.015** |
| Peanut, n (%) | 4 (28.6%) | 11 (36.7%) | 0.738 | 7 (43.8%) | 10 (30.3%) | 0.523 |
| Tree nut, n (%) | 4 (28.6%) | 16 (53.3%) | 0.195 | 10 (62.5%) | 11 (33.3%) | 0.070 |
| Storage proteins, n (%) | 5 (35.7%) | 15 (50.0%) | 0.519 | 9 (56.2%) | 15 (45.5%) | 0.551 |
| Milk, n (%) | 2 (14.3%) | 6 (20.0%) | 1.000 | 4 (25.0%) | 6 (18.2%) | 0.709 |
| HDM, n (%) | 4 (28.6%) | 9 (30.0%) | 1.000 | 5 (31.2%) | 8 (24.2%) | 0.733 |
| Birch pollen, n (%) | 5 (35.7%) | 10 (33.3%) | 1.000 | 5 (31.2%) | 11 (33.3%) | 1.000 |

Data presented as median (IQR) for continuous variables and n (%) for categorical variables. Clusters were derived independently by k-means clustering of skin tape strip (AD patients only, n = 44) and serum (n = 49) proteomic data. For comparison of clusters within each compartment, the Mann-Whitney U test was used for continuous variables and Fisher’s exact test for categorical variables. Statistically significant p-values (p < 0.05) are shown in bold. EASI: Eczema Area and Severity Index. oSCORAD: objective SCORAD. SCORAD: total SCORAD. IDQOL: Infants’ Dermatitis Quality of Life index. CDLQI: Children’s Dermatology Life Quality Index. TEWL: transepidermal water loss. NL: non-lesional. L: lesional. HDM: house dust mite.
